# Changes in foot progression angle during gait reduce the knee adduction moment and do not increase hip moments in individuals with knee osteoarthritis

**DOI:** 10.1101/2022.01.10.22268858

**Authors:** Kirsten Seagers, Scott D. Uhlrich, Julie A. Kolesar, Madeleine Berkson, Janelle M. Kaneda, Gary S. Beaupre, Scott L. Delp

## Abstract

People with knee osteoarthritis who adopt a modified foot progression angle (FPA) during gait often benefit from a reduction in the knee adduction moment. It is unknown, however, whether changes in the FPA increase hip moments, a surrogate measure of hip loading, which will increase the mechanical demand on the joint. This study examined how altering the FPA affects hip moments. Individuals with knee osteoarthritis walked on an instrumented treadmill with their baseline gait, 10° toe-in gait, and 10° toe-out gait. A musculoskeletal modeling package was used to compute joint moments from the experimental data. Fifty participants were selected from a larger study who reduced their peak knee adduction moment with a modified FPA. In this group, participants reduced the first peak of the knee adduction moment by 7.6% with 10° toe-in gait and reduced the second peak by 11.0% with 10° toe-out gait. Modifying the FPA reduced the early-stance hip abduction moment, at the time of peak hip contact force, by 4.3% ± 1.3% for 10° toe-in gait (p=0.005, d=0.49) and by 4.6% ± 1.1% for 10° toe-out gait (p<0.001, d=0.59) without increasing the flexion and internal rotation moments (p>0.15). Additionally, 74% of individuals reduced their total hip moment at time of peak hip contact force with a modified FPA. In summary, when adopting a FPA modification that reduced the knee adduction moment, participants, on average, did not increase surrogate measures of hip loading.

## Introduction

Of the 30 million Americans who suffer from osteoarthritis (OA) (Cisternas et al., 2016), an estimated 14 million have symptomatic knee OA (Deshpande et al., 2016). Mechanical loading, specifically joint forces, influence OA initiation and progression (Andriacchi et al., 2004; Felson, 2013). Since direct measurement of compressive joint forces in a native joint is not possible, joint moments, like the knee adduction moment (KAM), are often used as surrogate measures of joint loading. The KAM is correlated with knee OA presence (Baliunas et al., 2002), knee pain (Robbins et al., 2011; Thorp et al., 2007), knee cartilage degradation (Chehab et al., 2014), and predicts knee OA progression (Miyazaki, 2002). People can change their KAM by modifying features of their gait, such as trunk sway or walking speed (Hunt et al., 2008; Mündermann et al., 2004).

Gait modifications are an attractive intervention because they can reduce surrogate measures of knee loading, such as the KAM, without requiring surgery or external devices. One such gait modification is changing the foot progression angle (FPA), i.e., toeing-in or toeing-out. Altering the FPA requires changes to lower-body kinematics (Shull et al., 2013) and muscle coordination (Charlton et al., 2018; Rutherford et al., 2010). Toeing-in generally reduces the first peak of the KAM (Shull et al., 2013), whereas toeing-out reduces the second peak of the KAM (Guo et al., 2007). Although the clinical efficacy of prescribing the same FPA modification to all individuals remains unclear (Hunt et al., 2018), prescribing this intervention in a personalized manner can increase the achievable reductions in KAM (Uhlrich et al., 2018) and avoid cases where individuals increase their KAM. Felson et al., (2019) showed a personalized approach is effective for other KAM-reducing interventions. If personalized FPA retraining proves to be effective clinically, it could be deployed in the clinic with advancements in mobile sensors for estimating the KAM (Wang et al., 2020; Boswell et al., 2021) and providing biofeedback to change it (Xia et al., 2020).

As research on FPA modifications for people with knee OA progresses towards a deployable intervention, it is important to understand their effects more comprehensively, particularly at the hip. Studies that evaluate the efficacy of KAM-targeted gait modifications focus primarily on the effects at the knee, although there is a need to evaluate the effects of these modifications on other joints (Mündermann et al., 2005). People with knee OA have a 25% lifetime risk of developing hip OA (Murphy et al., 2010) due to shared risk factors, which place multiple joints at risk for OA (Felson et al., 2004; Issa et al., 2002). Although it is unknown how gait modifications will affect the hip joint, increased hip moments in all planes will increase mechanical demand on the soft tissues around the hip. Increased hip moments alter the compressive loading environment in the joint (Simic et al., 2011) and are associated with hip OA symptoms (Hall et al., 2019), presence (Maly et al., 2013), severity (Diamond et al., 2018), and progression (Tateuchi et al., 2017). The total knee joint moment, which is a summary measure of knee moments in all planes, has been used to evaluate gait modifications that have a multiplanar effect (Asay et al., 2018). Computing a similar measure at the hip (i.e., the total hip moment) may be a valuable way to estimate the net change in soft tissue demand at the hip from a gait modification for each individual. Some studies have examined the effect of FPA gait modifications on hip contact forces, kinetics, and kinematics to minimize torsional loading and loosening of total hip replacements (Bowsher and Vaughan, 1995) and to reduce hip contact forces for patients with hip pathology (Wesseling et al., 2015). Wesseling et al., (2015) found that hip kinematic perturbations that mimic clinical gait patterns, which could result in a modified FPA, reduced hip moments in a simulation study, but these results have not been verified experimentally. Toe-out gait, which increased the KAM in asymptomatic individuals, increased hip sagittal and frontal plane moments (Legrand et al., 2021); however, it is unclear whether these effects will be the same in individuals with knee OA who reduce their KAM with a modified FPA. Therefore, an important next step is to understand how interventions that target knee loading affect hip kinetics, kinematics, and spatiotemporal parameters of gait in an osteoarthritic population.

This study analyzes the biomechanical effects of a FPA modification on the hip in individuals with medial compartment knee OA who reduce their KAM with the modification. We hypothesize that changing the FPA will not increase hip moments or the total hip moment at time of peak hip contact force compared to baseline gait. Additionally, we performed an exploratory analysis to evaluate the effect of FPA on kinematics and spatiotemporal parameters. Our results provide insight into whether gait modifications for knee osteoarthritis increase mechanical demand at the hip.

## Methods

### Data Collection

Participant data were analyzed from a clinical study (clinicaltrials.gov NCT02767570) that trained people with medial compartment knee OA to walk with a modified FPA to reduce their peak KAM. One hundred seven people provided informed consent in agreement with a Stanford University Institutional Review Board protocol. Inclusion criteria included medial compartment knee OA grade between one and three on the Kellgren-Lawrence scale from anterior-posterior weightbearing radiographs; medial knee pain of three or greater on an 11-point numeric rating scale; the ability to walk safely on a treadmill without assistance for 25 minutes; and BMI less than 35. Participants were excluded if they had symptomatic arthritis in lower limb joints other than the knee that was more severe than their arthritic knee. The limb examined for each participant was chosen based on the presence of radiographic and symptomatic medial knee OA. If medial knee OA was present in both knees, the more symptomatic side was analyzed.

Experimental gait data were collected during a visit where different FPA modifications were evaluated for their effect on KAM peaks. Nineteen reflective markers were placed bilaterally on the 2^nd^ and 5^th^ metatarsal heads, calcanei and medial and lateral malleoli, medial and lateral femoral epicondyles, anterior and posterior superior iliac spines, and the C7 vertebrae, with eighteen additional markers for limb tracking. First, a standing calibration trial was performed, followed by a functional hip joint center calibration trial during which participants were asked to circumduct their hips (Piazza et al., 2004). Participants then walked naturally for two minutes (i.e., baseline trial) at a self-selected speed on an instrumented split belt treadmill (Bertec Corporation, Columbus, OH, USA) while ground reaction force data and optical marker positions were recorded (Motion Analysis Corporation, Santa Rosa, CA, USA). Then, two-minute FPA modification trials were performed where vibrotactile biofeedback (Engineering Acoustics, Casselberry, FL, USA) was given to participants to toe-in and toe-out by 10° relative to their natural FPA. The absolute FPA was calculated as the angle between the line connecting the calcaneus and second metatarsal head marker positions and the forward direction of the treadmill during 15-40% of stance (Rutherford et al., 2008).

Of the one hundred seven participants who completed this task in the clinical study, fifty were included in this analysis (Table 1). Selection for this analysis required participants to reduce the first or second peak KAM by at least 5%, a magnitude of KAM reduction that would likely elicit an improvement in knee pain (Erhart-Hledick et al., 2012), with either a 10° toe-in or a 10° toe-out gait modification as compared to baseline. Participants who did not reduce their peak KAM would not be prescribed a FPA modification and were therefore excluded from this analysis. The first and second peaks of the KAM during the baseline walking trial were defined as the maximum values during 0-50% and 51-100% of the stance phase, respectively. The FPA modification KAM peaks were identified within ±10% of the timing of the participant’s baseline peaks. Participants were excluded from this analysis if they used the treadmill handrails during walking trials or had greater than an estimated 1 cm of soft tissue between the anterior superior iliac spine marker and the anatomical bony landmark to ensure the fidelity of hip joint marker data (Hicks et al., 2015; Leardini et al., 2005).

**Table 1:** Demographics of the fifty participants with symptomatic medial knee osteoarthritis included in the analysis. Standard deviation (SD) is reported in parentheses.

| Characteristic (N = 50) | Mean (SD) |
| --- | --- |
| Age (yr) | 61 (7) |
| Height (m) | 1.7 (0.1) |
| Mass (kg) | 76.2 (15.4) |
| BMI (kg/m<sup>2</sup>) | 25.8 (3.8) |
| Gender | M: 31<br>F: 19 |
| Static Knee Angle (°):<br>Varus (-) Valgus (+) | Ipsilateral: -2.4 (3.6)<br>Contralateral: -1.0 (3.6) |
| Participants who reduced peak KAM with a<br>modified FPA | Peak 1 only: n =11<br>Peak 2 only: n =14<br>Peak 1 & Peak 2: n =25 |
| Gait Speed (m/s) | 1.2 (0.1) |
| Kellgren and Lawrence Grade | □: 12, □: 27, □: 11 |
| Medial Knee Pain:<br>0 (no pain) - 10 (worst pain) | 4 (2) |

### Musculoskeletal Modeling

We used a constrained musculoskeletal model (Rajagopal et al., 2016) with 25 degrees of freedom for kinematic and kinetic analysis. This model includes a three degree of freedom hip joint with Euler flexion, abduction, and rotation angles. We adapted the model published by Rajagopal and colleagues, by locking the metatarsophalangeal joints, removing the arms, and including a two degree of freedom knee joint (Lerner et al., 2015).

Scaling the generic musculoskeletal model to match the dimensions of each subject was a two-step process. First, long-leg radiograph measurements were used to adjust frontal plane lower limb alignment and pelvic width. The frontal plane lower limb alignment was found by measuring the angle (ImageJ, National Institute of Mental Health, Bethesda, Maryland, USA) between a line connecting the hip and knee joint centers and a line connecting the knee and ankle joint centers (Lerner et al., 2015). The pelvic width was defined as the distance between the hip joint centers identified on the radiograph. Second, the remainder of the model bodies were scaled using locations of joint centers and anatomical markers in OpenSim 4.0 (Seth et al., 2018).

Inverse kinematics was performed to compute hip adduction, flexion, and internal rotation angles. Results were filtered with a 4^th^ order Butterworth filter and cutoff frequency of 15 Hz. These kinematics and ground reaction forces, also filtered at 15 Hz, were used in inverse dynamics to calculate external hip abduction, extension, and internal rotation moments as well as the KAM. Moments were reported in the joint coordinate reference frame.

### Data and Statistical Analysis

For each gait trial (baseline, 10° toe-in, and 10° toe-out), the last 20 steps that achieved the target FPA, within 2.5°, were analyzed. Each biomechanical metric of interest was calculated for every step and averaged over each walking condition.

Hip moments were normalized by participant mass (Wesseling et al., 2015) and compared between walking conditions for the primary kinetic metrics: the early-stance hip moment, total hip moment, and angular impulse. To estimate the hip moment at the time of greatest hip loading, the early-stance hip moment was computed as the average moment during 15-20% of stance phase, the time of first peak hip contact force (Bergmann et al., 2001; Wesseling et al., 2015). To estimate the net effect of moment changes in all planes, the total hip moment at time of peak hip contact force was calculated. This metric was defined as the square root of the sum of squares of the early stance hip abduction, flexion, and internal rotation moments (Asay et al., 2018). For analysis during the entire stance phase, the angular impulse value was computed as the area under the absolute value of the hip moment curve during stance phase. For kinematic analysis, joint angles were averaged during stance and compared between walking conditions to capture global changes. Peak angles were also compared between conditions. Spatiotemporal parameters were also determined for comparison, including FPA, stride length, and step width. Stride length was defined as the difference in right and left foot calcaneus marker positions at time of heel strike. Step width was defined as the distance between the right and left foot center of pressure at 50% of stance (Donelan et al., 2001).

Statistical evaluations of 10° toe-in and 10° toe-out gait metrics compared to baseline were performed in MATLAB R2020a (MathWorks Corporation, Natick, MA USA) using a Wilcoxon signed-rank test (α = 0.05), since a Kolmogorov-Smirnov test indicated the data were not normally distributed. Bonferroni corrected p-values for multiple comparisons across FPA conditions were reported to mitigate type I error. Additionally, the Cohen’s d effect size was calculated for further comparison of the hip moments between gait patterns, where d<0.20 is a small effect and d>0.80 is a large effect.

## Results

Participants adopted a FPA modification that reduced the peak KAM and did not increase surrogate measures of hip loading. On average, 10° toe-in modifications reduced the first peak of the KAM by 7.6% compared to baseline, and 10° toe-out reduced the second peak by 11.0%. The early-stance hip abduction moment was significantly lower during 10° toe-in gait (-4.3% ± 1.3%, [range: -31.7%, 11.0%], p=0.005, d=0.49) and during 10° toe-out gait (-4.6% ± 1.1%, [range: - 21.9%, 9.4%], p<0.001, d=0.59) compared to baseline. There were no significant differences (p>0.05, d<0.25) in the early-stance hip extension moment or early-stance internal rotation moment for 10° toe-in or toe-out conditions compared to baseline (Figure 1). The baseline internal rotation moment and changes with FPA modifications were small in magnitude (Figure 2, Table 2), yet this results in large percent changes from baseline (Figure 1).

**Figure 1:**
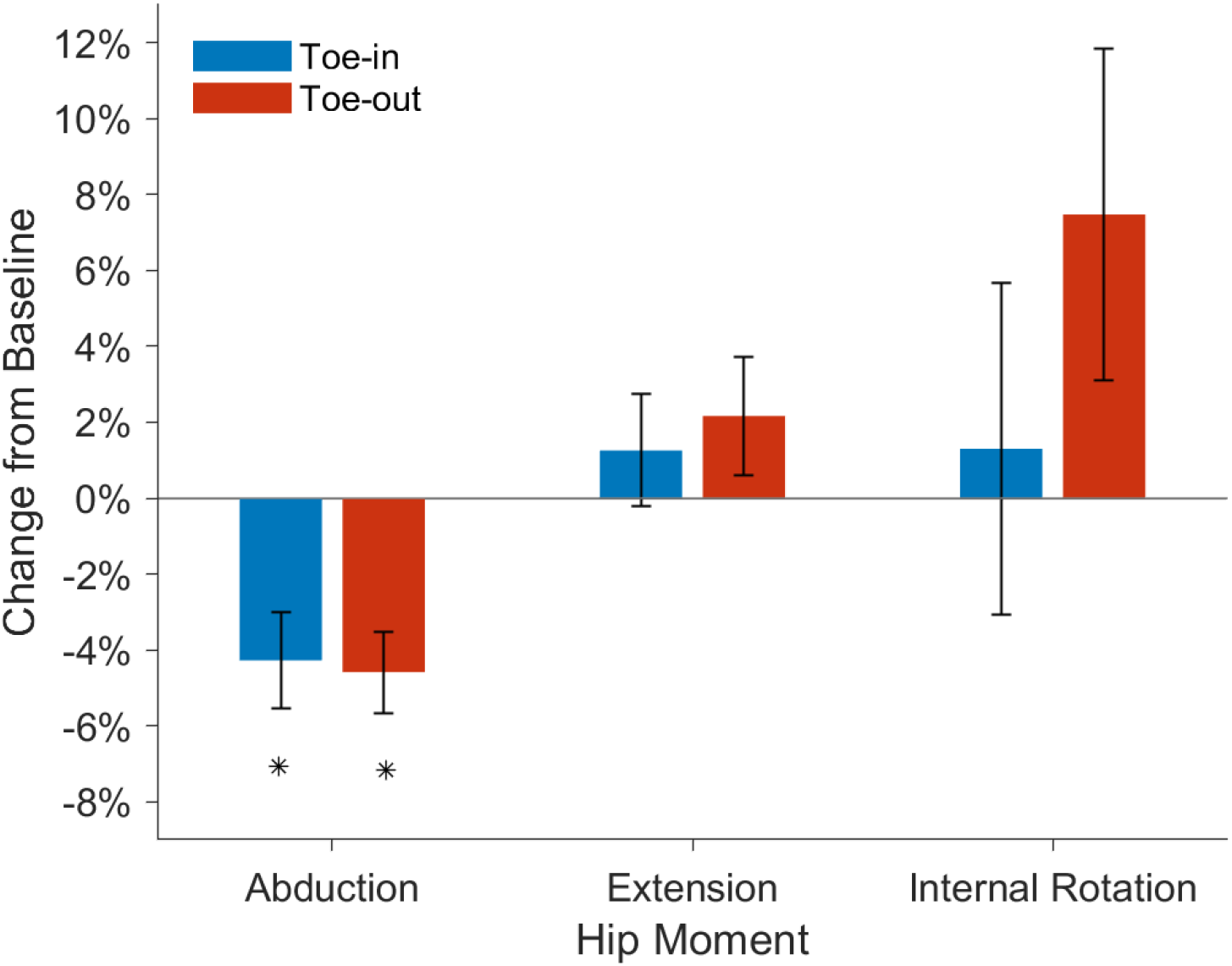
Participants reduced their early-stance hip abduction moment for 10° toe-in (blue) and 10° toe-out (red) gait as compared to baseline. No significant differences were observed between baseline gait and toe-in or toe-out gait for the early-stance hip extension and internal rotation moment. (*p<0.005)

**Figure 2:**
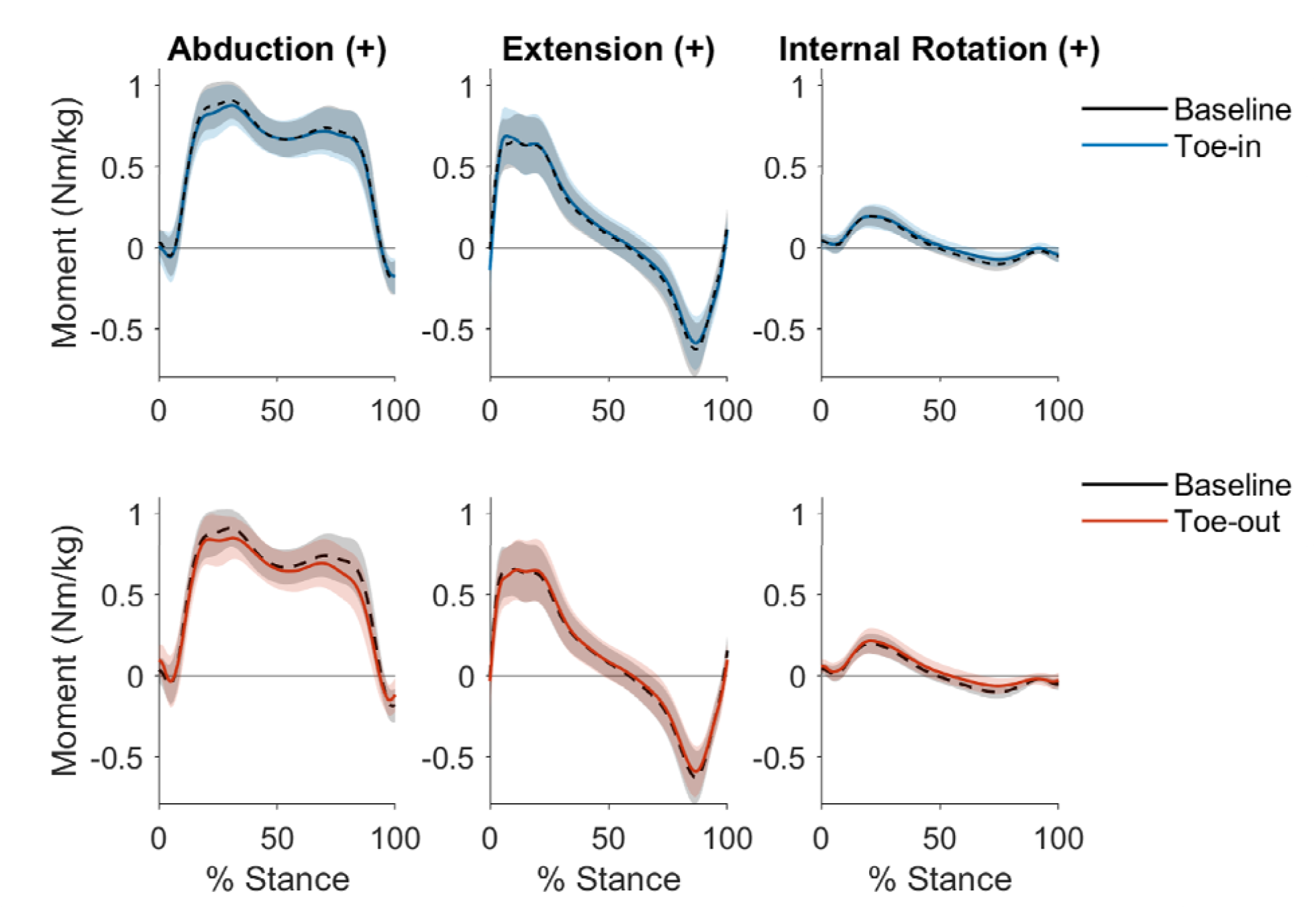
Ensemble-averaged hip moment curves, normalized by bodyweight, for all participants (n = 50) during stance phase. Baseline gait (black, dashed) hip moments are compared to 10° toe-in gait (blue, solid) in the first row and 10° toe-out gait (red, solid) in the second row. The shaded region indicates one standard deviation from the average moment.

**Table 2:** Spatiotemporal parameters, peak hip moments, and KAM for participants during baseline gait, 10° toe-in and 10° toe-out. Note, p-values are not reported for the peak KAM values since participants were selected based on their ability to reduce their peak KAM.

| Parameter | Baseline |  | 10° Toe-in |  |  | 10° Toe-out |  |  |
| --- | --- | --- | --- | --- | --- | --- | --- | --- |
|  | mean | SD | mean | SD | p-value | mean | SD | p-value |
| FPA (°) | 6.9 | 3.4 | - 2.8 | 3.5 | < **0.001** | 16.5 | 3.4 | < **0.001** |
| Stride Length (m) | 1.33 | 0.14 | 1.29 | 0.13 | < **0.001** | 1.29 | 0.13 | < **0.001** |
| Step Width (m) | 0.18 | 0.04 | 0.19 | 0.04 | 0.863 | 0.22 | 0.05 | < **0.001** |
| Peak Hip Abduction Moment (Nm/kg) | 0.97 | 0.12 | 0.93 | 0.13 | <**0.001** | 0.91 | 0.14 | <**0.001** |
| Peak Hip Extension Moment (Nm/kg) | 0.72 | 0.18 | 0.75 | 0.18 | **0.003** | 0.73 | 0.19 | 0.227 |
| Peak Hip Flexion Moment (Nm/kg) | -0.64 | 0.17 | -0.60 | 0.16 | <**0.001** | -0.60 | 0.15 | <**0.001** |
| Peak Hip Internal Rotation Moment (Nm/kg) | 0.20 | 0.06 | 0.21 | 0.08 | 0.37 | 0.22 | 0.08 | **0.025** |
| Peak Hip External Rotation Moment (Nm/kg) | -0.11 | 0.03 | -0.09 | 0.04 | <**0.001** | -0.08 | 0.04 | <**0.001** |
| 1st Peak KAM: (%BW*H) | 3.69 | 1.17 | 3.41 | 1.11 |  | 3.80 | 1.23 |  |
| (Nm/kg) | 0.62 | 0.20 | 0.57 | 0.19 |  | 0.64 | 0.21 |  |
| 2nd Peak KAM: (%BW*H) | 2.72 | 0.98 | 2.72 | 1.01 |  | 2.42 | 1.06 |  |
| (Nm/kg) | 0.46 | 0.17 | 0.46 | 0.17 |  | 0.41 | 0.18 |  |

On average, there were no significant increases in the total hip moment at time of peak hip contact force; however, on an individual basis when walking with a modified FPA that maximally reduced their KAM, 37 individuals (74% of participants) reduced their total hip moment, while 13 individuals (26% of participants) increased their total hip moment. When walking with a toe-out gait, 46% of individuals increased this metric, and 40% increased it with toe-in gait. The largest observed total hip moment increase from baseline was 9.2% and 12.4% for 10° toe-in and 10° toe-out gait, respectively. There was no difference in the change in total hip moment from baseline for 10° toe-in gait (-1.9% ± 0.8%, [range: -17.3%, 9.2%], p>0.05, d=0.30) or 10° toe-out gait (-1.4% ± 0.8%, [range: -16.5%, 12.4%], p>0.05, d=0.22) compared to baseline.

The frontal plane angular impulse was significantly reduced compared to baseline for 10° toe-in gait (-2.3% ± 0.7%, [range: -11.7%, 7.4%], p<0.01, d=0.45) and for 10° toe-out gait (-6.9% ± 0.8%, [range: -23.1%, 4.5%], p<0.001, d=1.23), but no significant differences (p>0.05, d<0.18) were observed in the sagittal or transverse plane (Figure 3).

**Figure 3:**
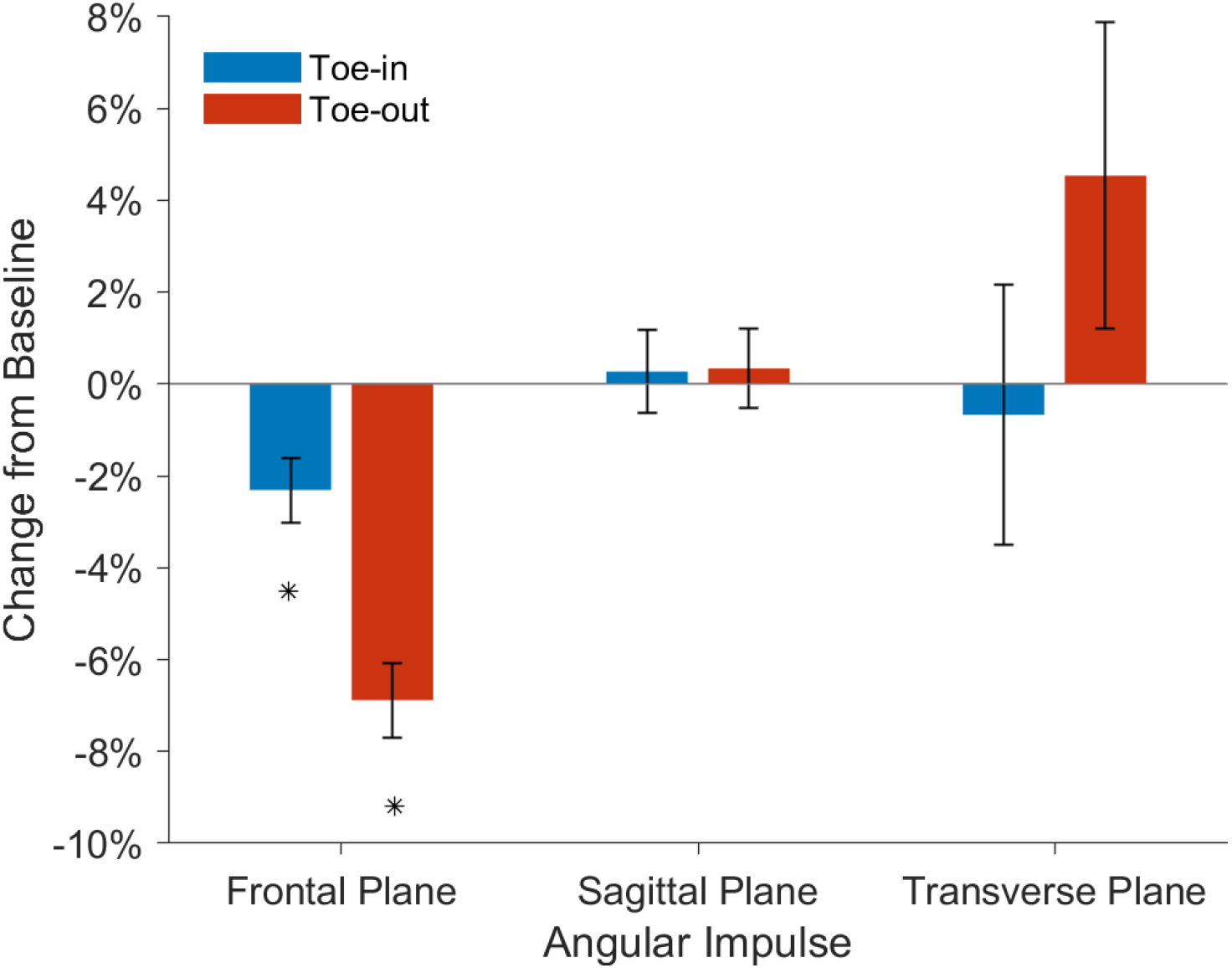
Participants reduced their frontal plane angular impulse during 10° toe-in (blue) and 10° toe-out (red) gait compared to baseline. No significant differences were observed between baseline and toe-in or toe-out gait for the angular impulse in the sagittal and transverse plane. (*p<0.01)

Changes in hip kinematics were minimal. Participants walked with a decreased *average* hip adduction angle for 10° toe-in gait (-0.4° ± 1.1°, p=0.045) and 10° toe-out gait (-1.6° ± 1.2°, p<0.001) compared to baseline (Figure 4). *Peak* hip adduction angles also decreased for both toe-in (-0.7° ± 1.5°, p=0.009) and toe-out (-1.8° ± 1.5°, p=0.005) gait compared to baseline. Participants walked with increased *average* hip flexion when toeing-in (2.7° ± 2.2°, p<0.001) and toeing-out (1.7° ± 2.1°, p<0.001). Both modifications had a lower *peak* extension angle compared to baseline (toe-in: -3.2° ± 2.7°, p<0.001; toe-out: -2.5° ± 2.7°, p<0.001). The *average* hip internal rotation angle increased by an average of 4.4° ± 2.6° (p<0.001) when walking with a 10° toe-in gait and decreased by 5.7° ± 1.3° (p<0.001) with a 10° toe-out gait. Similar directional changes were seen in the *peak* hip internal rotation angle compared to baseline (toe-in: 4.6° ± 2.7°, p<0.001; toe-out: -5.8° ± 1.6°, p<0.001). Changes in spatiotemporal parameters were also observed. Stride length decreased with both FPA modifications (p<0.001), while step width increased only for 10° toe-out gait (p<0.001) (Table 2).

**Figure 4:**
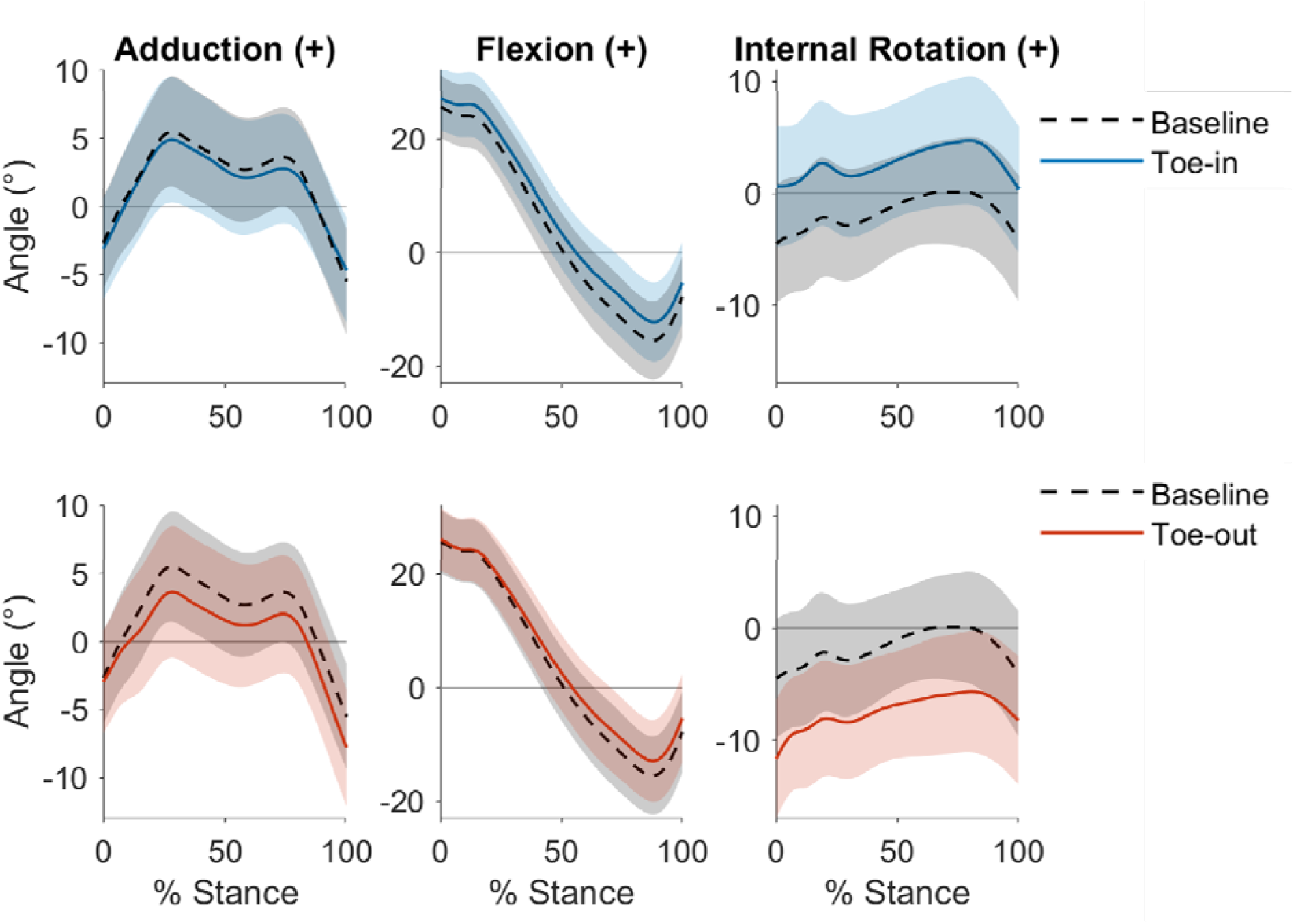
Ensemble-averaged hip kinematics for all participants (n = 50) during stance phase. Baseline gait (black, dashed) hip angles are compared to 10° toe-in gait (blue, solid) in the first row and 10° toe-out gait (red, solid) in the second row. The shaded region indicates one standard deviation from the average hip angle. The peak hip adduction angle and the peak hip extension angle are significantly reduced with 10° toe-in gait and 10° toe-out gait compared to baseline (p<0.01). The peak hip internal rotation angle is significantly increased for 10° toe-in gait and reduced for 10° toe-out gait compared to baseline (p<0.001).

## Discussion

We analyzed how changing the FPA during walking affects hip moments in fifty individuals with medial knee OA who successfully reduce their KAM with the modification. Our hypothesis was confirmed, as hip moments did not increase, on average, when a modified FPA was adopted. Both 10° toe-in and 10° toe-out modifications decreased the early-stance hip abduction moment and angular impulse. On average, adopting a FPA modification that reduced the KAM did not significantly increase hip moments at time of peak hip contact force, the total hip moment at time of peak hip contact force, or the angular impulse. However, some participants did increase their total hip moment at time of peak hip contact force with the FPA modification. This suggests that for individuals with hip joint pathology or at risk of developing hip OA, the effect of a FPA gait modification at the hip should be evaluated individually to ensure that adopting a modification that improves knee loading does not cause a potentially harmful increase in hip loading.

Our observed decrease in the hip abduction moment is consistent with other studies. The 4% decrease in the early-stance hip abduction moment when toeing-in and toeing-out was smaller than the 16% decrease in hip abduction observed at the time of peak KAM in a healthy population (Kettlety et al., 2018). This could be due to the varying participant population and differences in gait cycle timing when the hip moments were analyzed (i.e., at peak hip contact force versus peak KAM). Similarly, the reductions in early-stance hip abduction moment (10° toe-in: 0.03 Nm/kg; 10° toe-out: 0.04 Nm/kg) agreed directionally with the 0.1 Nm/kg decrease Wesseling et al., (2015) reported from simulated perturbations of frontal plane hip kinematics. The magnitude differences are unsurprising when comparing simulation predictions to experimental results, since participants may adopt other compensatory strategies to modify their FPA that were not captured in these simulations. Additionally, our observed hip rotation moment changes were small in magnitude, compared to the abduction and extension moments, resulting in large percent changes.

Kinematic and spatiotemporal changes were also observed when participants modified their FPA. Changes in the hip rotation angle were less than the change in FPA, which aligns with results from Cibulka et al., (2016), who found that FPA modifications elicit changes at multiple lower-extremity joints. In addition to the rotational kinematics changes, the average hip adduction angle decrease relates to the increase in step width for toe-out gait. Other studies (Kettlety et al., 2020; Uhlrich et al., 2018) have also observed that step width increases with a modified FPA resulting from less hip adduction. While the decrease in hip adduction angle and increase in step width are small, these changes increase medio-lateral stability (Meyer et al., 2018). Additionally, participants decreased stride length, which can increase double-limb support time, resulting in a more stable gait (Shkuratova et al., 2004). The changes to kinematics and spatiotemporal parameters may attenuate over time as an individual becomes more comfortable with the gait modification.

It is important to identify the limitations of our study. We estimated the effect of FPA modifications on surrogate measures of hip loading. Electromyography was not used in this study, but, in the future, validated electromyography-informed simulations could estimate changes in muscle and hip contact forces. The surrogate measure of hip loading that we used, joint moments, is related to muscle and joint contact forces, which provides an estimate of the directional change in hip contact force. Meyer et al. (2018) reported that a decrease in the hip abduction moment and hip adduction angle reduced the work generated by the hip abductor muscles and decreased hip loading in people with hip OA. The relationship between the total hip moment and hip OA pathogenesis has not been studied, but the total knee joint moment and the change in relative contribution of its components (knee adduction, flexion, and rotation moments) reflect gait changes associated with progressing knee OA and pain (Asay et al., 2018). Similarly, the total hip moment is likely an effective summary metric of changes in muscular demand in all planes at the hip and future work should investigate how it relates to hip OA initiation and progression. Generally, little is known about the role of hip joint loading on the initiation of hip OA; elucidating these relationships should be the topic of future work.

Several studies have shown that FPA modifications can provide meaningful reductions in the KAM. Our results add to this growing body of evidence and show that, on average, FPA modifications reduce frontal plane hip moments without increasing the moments in other planes. However, some individuals did increase their total hip moment at time of peak hip contact force. If an individual is at risk for hip OA, then hip moments will be an important biomechanical parameter to monitor when prescribing gait modifications. Yet, 74% of people, who could meaningfully reduce their KAM, reduced their total hip moment at time of peak hip contact force with a modified FPA. In summary, on average and for most individuals, a FPA modification that reduces knee loading does not increase surrogate measures of hip loading.

## Data Availability

All data produced in the present study are available upon reasonable request to the authors once the paper is published in a peer reviewed journal.

## Acknowledgements

We would like to thank the participants involved in the study. This work was supported by the Sang Samuel Wang Stanford Graduate Fellowship, NSF Graduate Research Fellowships (DGE-1147470 and DGE-1656518), NIH grant P41EB027060, and by Merit Review Award I01 RX001811 from the United States Department of Veterans Affairs Rehabilitation Research and Development Service.

